# Development of an artificial intelligence-generated, explainable treatment recommendation system for urothelial carcinoma and renal cell carcinoma to support multidisciplinary cancer conferences

**DOI:** 10.1101/2025.02.03.25321592

**Authors:** Gregor Duwe, Dominique Mercier, Verena Kauth, Kerstin Moench, Vikas Rajashekar, Markus Junker, Andreas Dengel, Axel Haferkamp, Thomas Höfner

## Abstract

**Background and Objective:** Decisions on the best available treatment in clinical oncology are based on expert opinions in multidisciplinary cancer conferences (MCC). Artificial intelligence (AI) could increase evidence-based treatment by generating additional treatment recommendations (TR). We aimed to develop such an AI system for urothelial carcinoma (UC) and renal cell carcinoma (RCC).

**Methods:** Comprehensive data of patients with histologically confirmed UC and RCC who received MCC recommendations in the years 2015 – 2022 were transformed into machine readable representations. Development of a two-step process to train a classifier to mimic TR. Identification of superordinate categories of recommendations followed by specification of detailed TR. Machine learning (CatBoost, XGBoost, Random Forest) and deep learning (TabPFN, TabNet, SoftOrdering CNN, FCN) techniques were trained. Results were measured by F1-scores for accuracy weights. Additionally, clinical trial data for drugs were included.

**Key Findings and Limitations:** AI training was performed with 1617 (UC) and 880 (RCC) MCC recommendations (77 and 76 patient input parameters). AI system generated fully automated TR with excellent F1-scores for UC (e.g. ‘Surgery’ 0.81, ‘Anti-cancer drug’ 0.83, ‘Gemcitabine/Cisplatin’ 0.88) and RCC (e.g. ‘Anti-cancer drug’ 0.92 ‘Nivolumab’ 0.78, ‘Pembrolizumab/Axitinib’ 0.89). Explainability is provided by clinical features and their importance score. TR and explainability were visualized on a dashboard. Main limitations: single-centre and retrospective study.

**Conclusions and Clinical Implications:** First AI-generated explainable TR in UC and RCC with excellent performance results. Potential support tool for high-quality, evidence-based TR in MCC. Study sets global reference standards for AI development in MCC recommendations in clinical oncology.

**Patient Summary:** Our artificial intelligence tool is designed to help experts at multidisciplinary cancer conferences make personalized treatment recommendations for each patient with urothelial or renal cell cancer. It was trained with extensive specific patient data and provides explanations of how recommendations were generated with good outcomes. Artificial intelligence can lead to improved treatment recommendations by providing a more comprehensive view of the individual patient in multidisciplinary decision making.

## Introduction

Worldwide, genitourinary (GU) malignancies represent approximately 26% of all new cancer diagnoses^1^. Finding the best treatment for patients with advanced cancer is a complex task, including expert knowledge of numerous medical disciplines^2^. The primary objective is to provide the optimal treatment for the patient, taking into account the probability of success, potential risks of pharmaceuticals and surgical procedures, as well as the patient’s life expectancy. These critical treatment decisions are made in multidisciplinary cancer conferences (MCC)^3^. The emerging complexity in clinical oncology has made these decisions more difficult as new drugs frequently modify the treatment^4,5^. Given the considerable volume of information to be considered and the unique clinical characteristics of individual patients, there is a risk for experts in MCC to overlook developments specific to the patient’s case, or to misidentify risks and side effects.

Artificial intelligence (AI) has the potential to reduce this complexity for physicians while increasing the level of evidence-based treatment recommendations (TR)^6,7^. AI in clinical oncology has primarily been explored for the prediction of survival rates and responses of individual treatments^8–11^ as well as the processing of radiological and pathological imaging data with excellent outcome data^10^. However, text data is processed less frequently and only in a few cases TR are subsequently provided^12,13^. With our multidisciplinary project (ger. “KITTU”: AI-supported Clinical Decision in Urologic Oncology), we aim to support physicians in MCC by generating individual, explainable evidence-based AI-generated TR for urothelial carcinoma (UC) and renal cell carcinoma (RCC) patients^6^. Furthermore, such an AI system is ideally suited to the increasing digital transformation of our healthcare systems by reducing workload while providing high quality decision support.

## Materials and Methods

### Patient cohort, data security and data preprocessing for network training

All patients ≥18 years with histologically confirmed UC and RCC who received MCC recommendations at the University Medical Center Mainz (UMCM) in the years 2015- 2022 were screened for study inclusion. Ethical and legal frameworks are shown in Supplement S1.1. Cases not suitable for AI training were excluded for reasons shown in Figure 1.

**Fig. 1:**
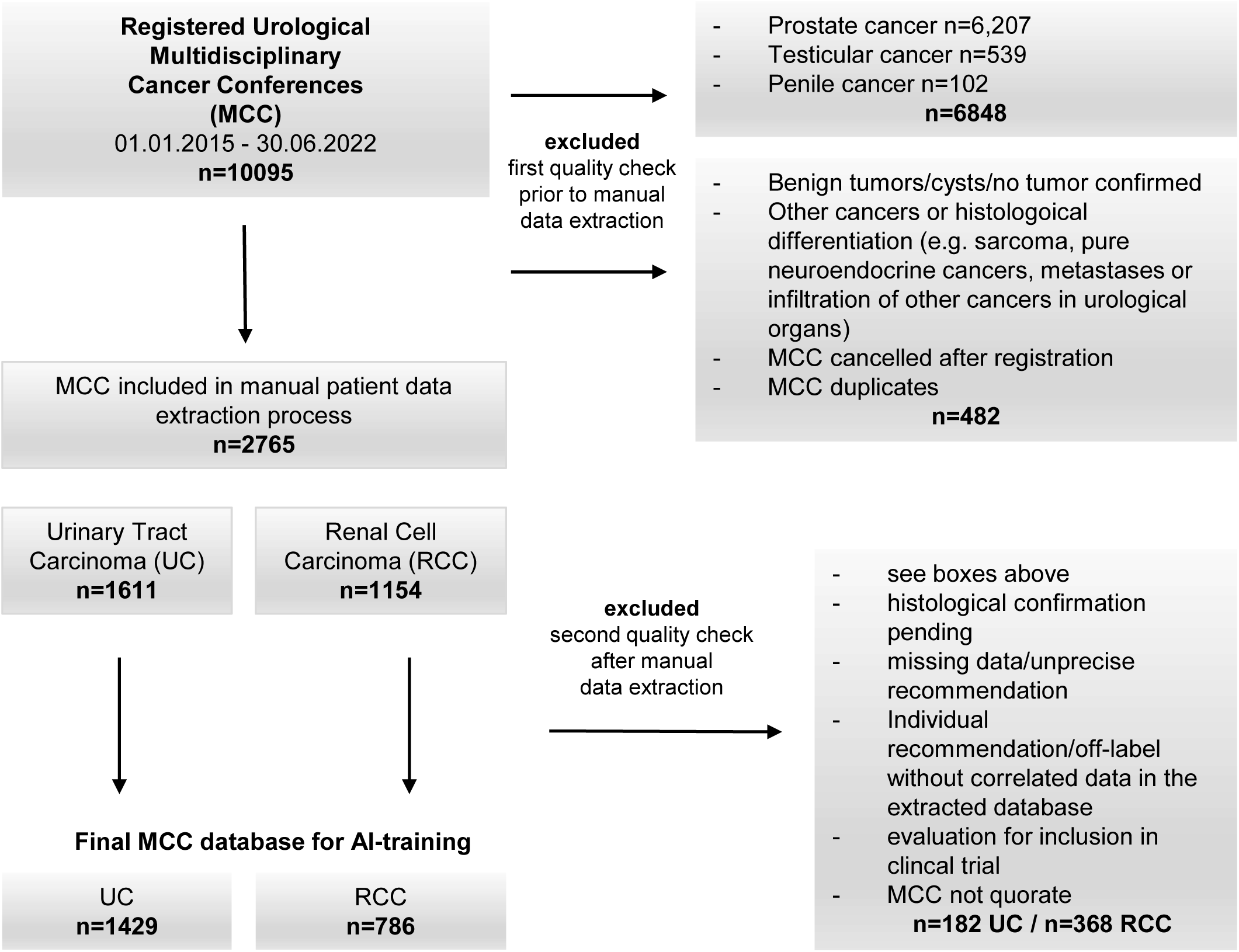
Flow chart of included and excluded MCC cases for AI training.

Retrospective patient data was included in this pre-clinical study for the initial technical development of AI software. MCC documentation, including TR as well as the corresponding extensive patient data, was extracted manually from the UMCM SAP system by medical professionals, followed by manual quality control. Complete clinical data structure is listed in Supplement Tab. 1.

Pseudonymised data were transferred to the German Center for Artificial Intelligence (DFKI) via a secure, locally hosted cloud storage and further normalized and standardized for machine learning architectures (see Supplement S1.2./S1.3.). Since it was not possible to directly train a network for multi label prediction due to the low number of instances, a counter intuitive decision was established for MCCs with multiple equivalent recommendations by means of duplicating them to achieve respective single recommendations (see Supplement S1.4.).

### High- and Low-level treatment recommendations

A significant technical advancement was the division of TR into High-level (superordinate, general class of recommendation, e.g. ‘Surgery’, ‘Anti-cancer drug’) and Low-level recommendations (specific recommendation, e.g. ‘Cystectomy’, ‘Pembrolizumab’). The large number of different drugs and surgical treatments (some with low case numbers) results in a high complexity which was tackled by the “divide and conquer principle” using a high- and low-level separation (Figure 2).

**Fig. 2:**
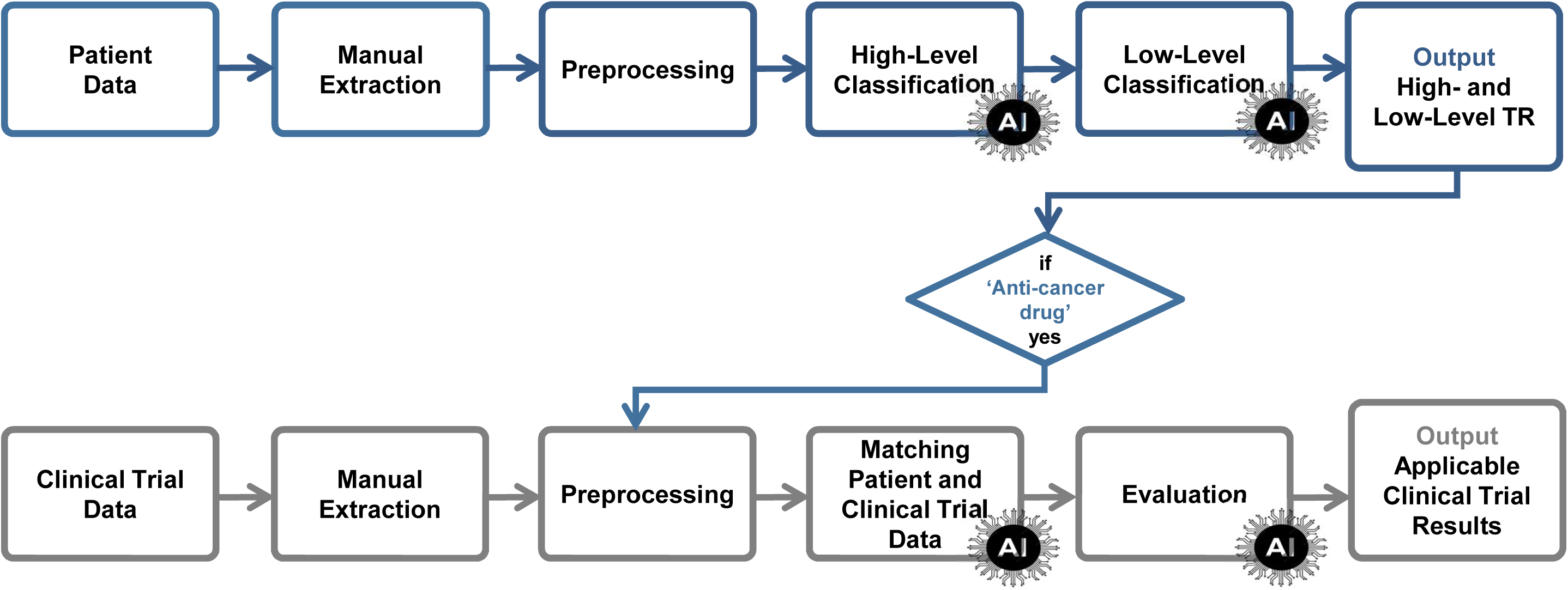
Workflow of data processing (detailed description in Supplement S1.5.)

### Performance evaluation

The data set with MCC recommendations was divided into a training set (90%) and a test set (10%). In the first step of AI training, High-level categories of recommendations were identified (Figures 3 and 4). In the second step, the specific recommendation (High-level and Low-level) is provided for each MCC case of the training set. Various classifiers were used: machine learning approaches (CatBoost, XGBoost, Random Forest) and deep learning algorithms (TabPFN, TabNet, SoftOrdering CNN, Fully Connected Networks).

**Fig.3:**
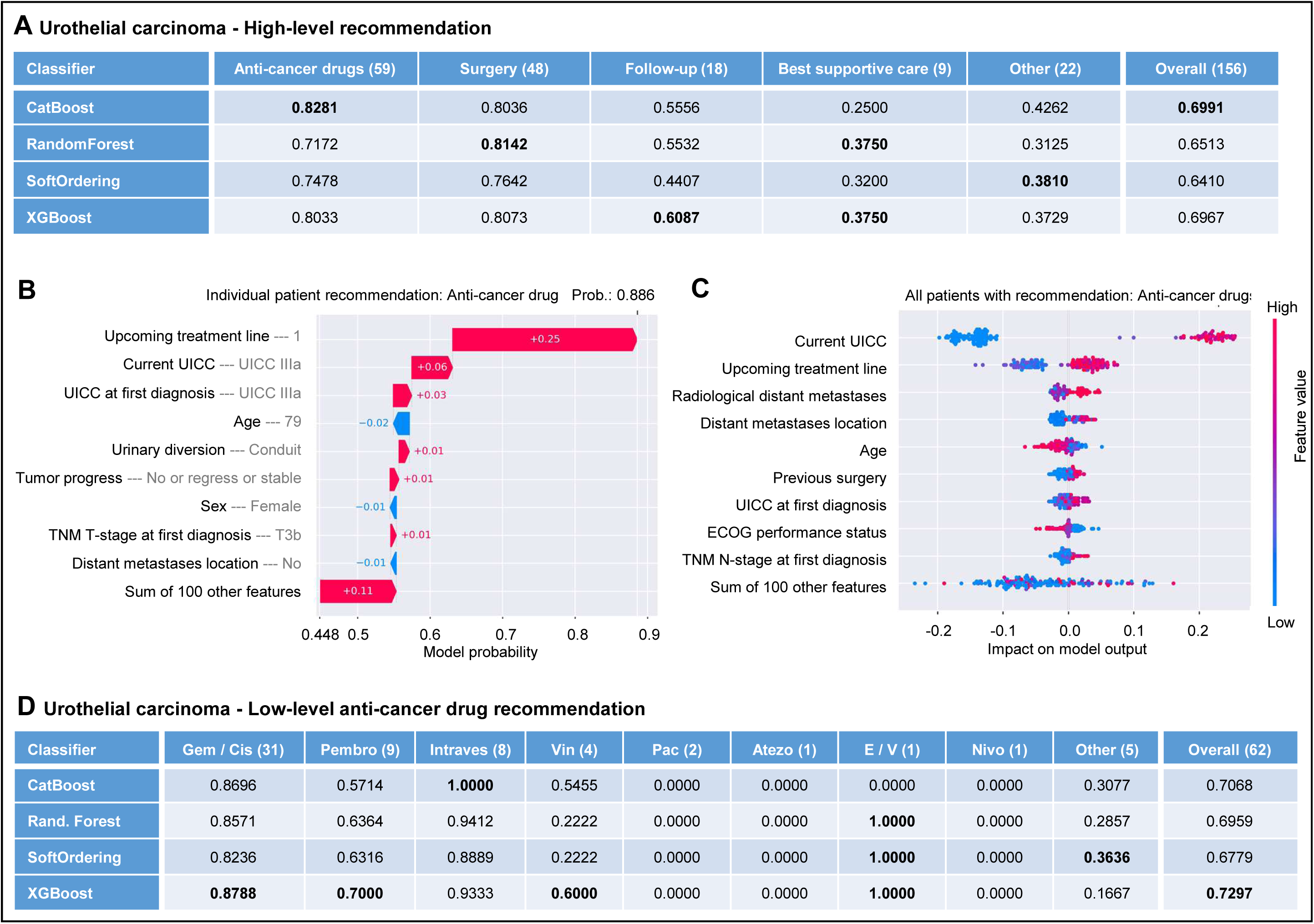
Urothelial carcinoma: AI performance results (test set) of different classifiers for High-level TR (A), Low-level ‘Anti-cancer drug’ TR (D) and Waterfall/Beeswarm plot for exemplary High-Level TR explainability (B+C) **A+D:** Numbers in brackets correspond to number of cases in the test set. Numbers in the table show the specific F1-scores. The ‘Other’ class always corresponds to a group of classes that is under-represented in the dataset and can be more specified in future once the dataset has a sufficient number of cases for reasonable AI training. **D:** Gem/Cis=Gemcitabin/Platin, Pembro=Pembrolizumab, Intraves=Intravesical, Vin=Vinflunin, Pac=Paclitaxel, Atezo=Atezolizumab, E/V=Enfortumab-Vedotin, Nivo=Nivolumab **Please note:** Total numbers of ‘Anti-cancer drug’ cases vary between A and D: The High-level AI (A) separated the classes into the 90% training and 10% test set and ensured the correct ratio of each class in both sets (the specific ratio for single drugs is not relevant here). The Low-level AI (D) separated the two sets focused on the specific drug to ensure the correct ratio of the single drug (e.g. Gem/Cis) in both sets which explains that more cases are found in the test set due to rounding up/off. **B:** Patient values in grey next to the feature names. Numbers In/next to the arrows = degree they contribute to the probability. **C:** Listed features are the ones that are most important to decide for or against the recommendation, impact is shown on the x- axis. Positive values favor the recommendation and negative favor a different recommendation. Distribution of the values in the dataset is shown in a point cloud. Red dots correspond to a high feature value (e.g. UICC IV) and blue dots to a low feature value (e.g. upcoming treatment line “firstline”).

**Fig.4:**
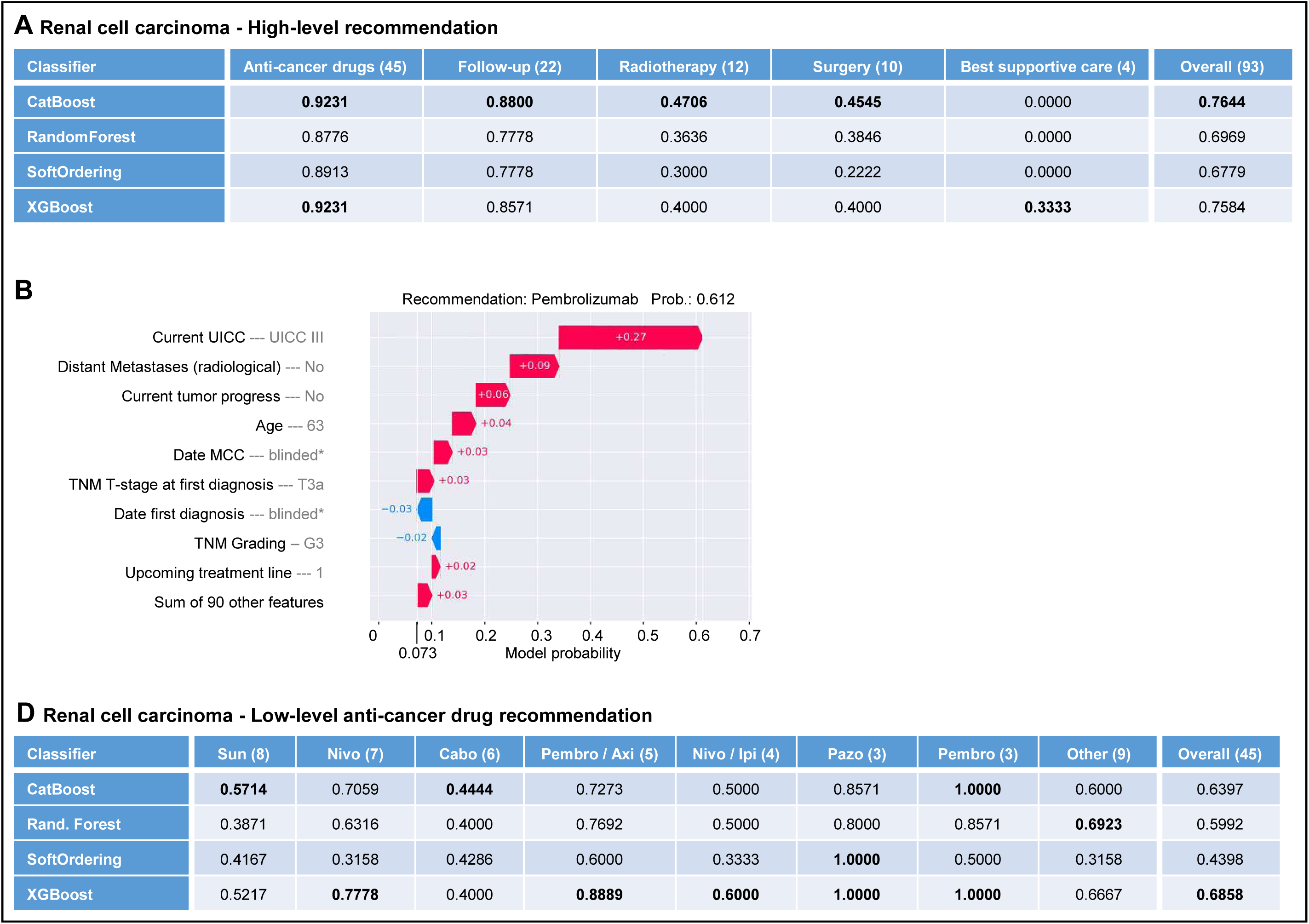
Renal cell carcinoma: AI performance results (test set) of different classifiers for High-level TR (A), Low-level ‘Anti-cancer drug’ TR (C) and Waterfall plot for exemplary Low-Level Pembrolizumab TR explainability (B) **A+C:** Numbers in brackets correspond to number of cases in the test set. Numbers in the table show the specific F1- scores. **C:** Sun=Sunitinib, Nivo=Nivolumab, Cabo=Cabozantinib, Pembro/Axi=Pembrolizumab/Axitinib, Nivo/Ipi=Nivolumab/Ipilimumab, Pazo=Pazopanib, Pembro=Pembrolizumab. **B:** Patient values in grey next to the feature names. Numbers in/next to the arrows = degree they contribute to the probability. *blinded is used to conceal the patient value in this manuscript for data protection, the original data set contains the real value which is taken into account by the AI.

For the test set, the accuracy of the AI-generated recommendations was determined by F1-scores. The F1 score is a machine learning evaluation metric that is calculated as the harmonic mean of precision and recall, ranging from 0 to 1, comparable to accuracy. It balances precision and recall, ensuring neither is ignored and performs well on imbalanced datasets where accuracy alone can be misleading. A higher F1 score indicates better overall model performance. Low-level categories with underrepresented, low case numbers (e.g. rarely recommended drug treatments) have been grouped into an ‘Other’ category to reduce untrainable categories. These TR can be differentiated in the future once a sufficient number of cases are available.

### Explainability based on patient characteristics and clinical trials

SHAP (SHapley Additive exPlanations) explanations^14^ for High- and Low-level recommendations highlight which patient feature contribute positively and negatively to a given recommendation by perturbing values and calculating their impact on the AI-generated recommendation. In addition, global explanations were created for the recommendation classes to identify the most relevant features across the total patient cohort. To enhance the recommendation and explainability, survival data of clinical trials for anti-cancer drugs were taken into account (Supplement S1.6.).

### Dashboard visualization

The MCC recommendation generated was visualized in a dashboard for the medical staff (Figure 5), including interactive features and clinical trial data to provide explanation for the recommended treatment.

**Fig.5:**
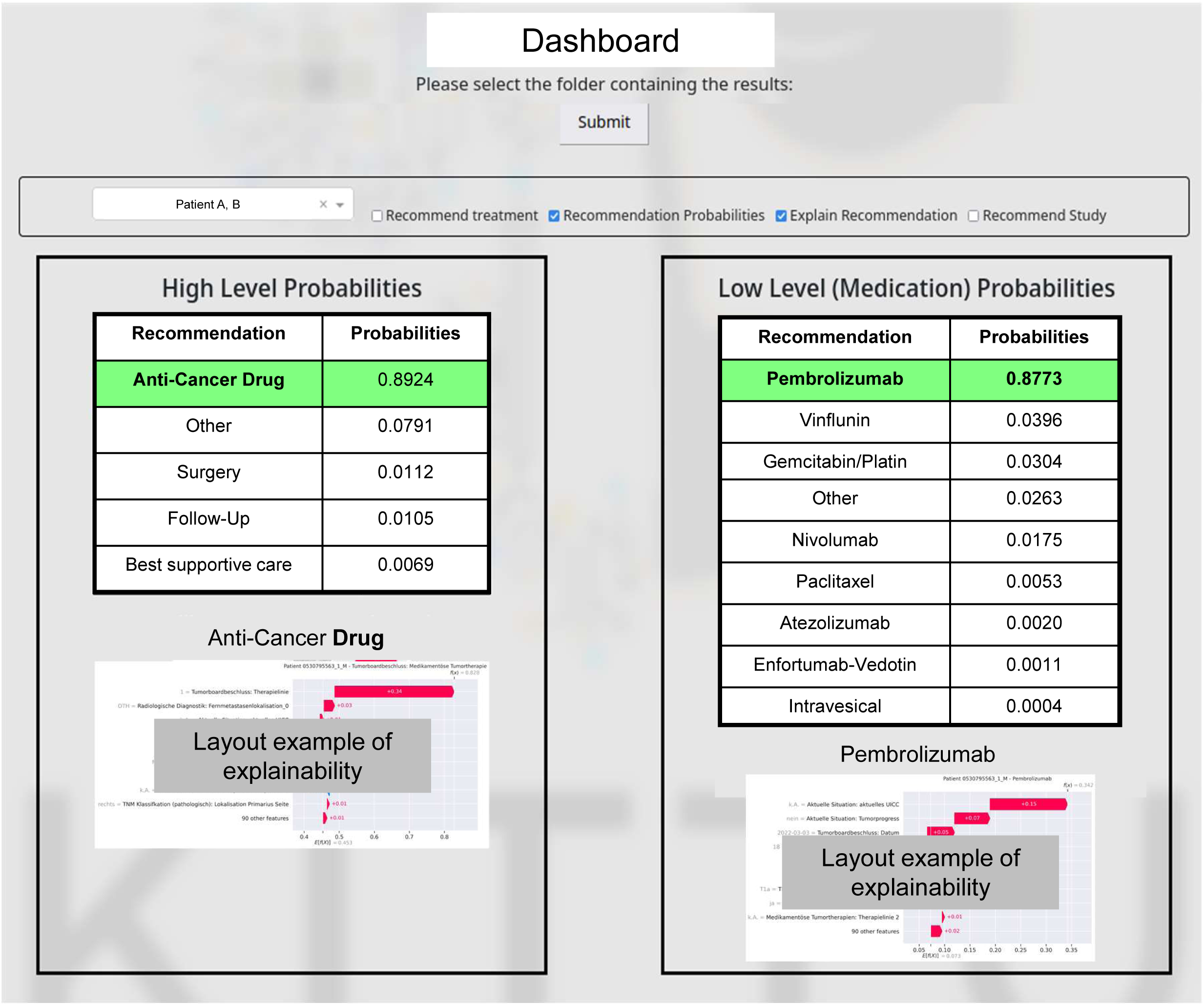
Exemplary dashboard for an individual UC patient. On the top left, the patient can be selected. Next, any components required can be selected to be shown. The High-level and Low-level TR are presented based on the probabilities with green-highlighted for the most emphasized AI-generated TR. Explainability graphs below.

## Results

### Artificial-Intelligence generated treatment recommendation for urothelial carcinoma

1429 MCC cases were included in the AI training for UC patients. After appropriate duplication of individual cases with more than one (equivalent) recommendation, the resulting total of 1617 cases were randomly divided into training and test sets (1461 and 156 cases). 77 individual patient input parameters were considered for each patient. Some features were distributed into machine-readable subfeatures, resulting in a final number of 109 employed patient characteristics used. Thirteen output parameters were added to enable assessment of the AI-generated TR (Supplement Tab. S1). The final results of AI-generated TR for UC are shown in Figure 3 (A+D). Overall F1-score for the best High-level performing model (CatBoost) achieved 0.6991. High-level ‘Surgery’ (0.8142) and ‘Anti-cancer drug’ (0.8281) recommendations yielded highest F1-scores. F1 scores for ‘Best supportive care’, ‘Follow-up’ and ‘Other’ showed lower F1 scores presumably due to the currently low caseload. For Low-level drug TR, the best classifier was XGBoost (overall F1 score 0.7297) with best performance for the classes Gemcitabin/Platin (0.8788) and Pembrolizumab (0.7000). Performance results for Low-level surgery recommendations are shown in Supplement S2.1.

### Artificial-Intelligence generated treatment recommendation for renal cell carcinoma

786 MCC cases were included in the AI training for RCC. After appropriate duplication of individual cases with more than one (equivalent) recommendation, the resulting total of 880 cases was randomly distributed to training and test set (787 and 93 cases). 76 individual patient input parameters were considered for each patient. Some features were divided into machine-readable subfeatures which resulted in a final count of 99 employed patient characteristics used. Eleven output parameters were included to enable assessment of the AI-generated TR (Supplement Tab. S1). The final results of AI-generated TR for RCC are shown in Figure 4 (A+C). The overall F1-score for the best High-level performing model (CatBoost) achieved 0.7644. The best results were obtained for High-level ‘Anti-cancer drug’ (0.9231) and ‘Follow-up’ (0.8800). For Low-level anti-cancer drug recommendations, XGBoost showed the best overall F1 result (0.6858) with best scores achieved for Pazopanib (1.000) and Pembrolizumab (1.000) (for assessment of F1-score 1.000 (see Supplement S2.1.) Performance results for Low-level surgery recommendations are shown in Supplement S2.2.

### Explainability and Dashboard Presentation

For the explainability of the AI-generated TR, an attribution method was used to identify the most relevant features during the computation of the recommendation. The SHAP model^14^ was used to generate an explanation for the individual patient recommendation (Figure 3B) and for a recommended class in general, taking into account all affected patients (Figure 3C) was generated. While the explanation of the patient recommendation shows the extent to which each feature and the corresponding patient value influenced the prediction positively or negatively. While the explanation of the patient recommendation shows the which extent to which each feature and the corresponding patient value affected the prediction positively or negatively, the visualization of the classes in general shows which values contributed most commonly to a recommendation.

In Figure 3B, an example case/ explanation shows a “probability” (Softmax) of 0.886. This describes the likelihood of the High-level ‘Anti-cancer drug’ recommendation by the AI based on the given recommendations in the training set (with 1.0 being the highest possible value). The “beeswarm explanation” in Figure 3C visualizes how different feature values affect the recommendation in the total dataset of all patients with this TR, with low values representing a negative influence and vice versa. The statistical graphs highlight which features can be assessed to vote for or against a TR based on the distribution of the dataset. The example shows that current UICC, upcoming treatment line and age were the main features that impacted the recommendation ‘Anti-cancer drug’.

The developed dashboard facilitates the practical use of the system in a clinical setting (Figure 5). It covers all relevant information of the TR, including the visualization of the explainability based on patient characteristics and clinical trial information in order to better understand and individually assess the AI-based TR (see Supplement S2.3.).

## Discussion

To our knowledge, we have developed the first AI assistance tool in clinical oncology to generate explainable MCC recommendations for UC and RCC with promising initial results. In the long term, it could enhance the quality of MCC worldwide. The developed AI algorithms are characterized by adherence to methodological standards, including the use of high quality patient data obtained through comprehensive manual extraction, the inclusion of a large number of clinical variables (“features”), and the formulation of recommendations at different TR levels (High-level/Low-level recommendation).

The decision to integrate multiple and unbiased clinical parameters in the development of the AI is based on the assumption that we cannot decide exactly which factors contribute to the relevant clinical decision for the patients from a routine AI perspective. Our AI system provides TR in urological oncology with a high degree of accuracy and additional explainability based on clinical features. The applicability of the AI-generated TR in daily practice is further enhanced by an interactive dashboard that can be used by the medical staff. Final treatment decisions will always be made by humans.

### Comparison of Artificial Intelligence Models of treatment recommendations in genitourinary oncology

Compared to other AI systems in Clinical Oncology, our AI routine was developed with complex tabular data which precluded the use of pre-trained models and required the implementation of extensive feature preprocessing. This successful approach has not been previously described. Previous GU cancer studies for AI-generated TR focused on localized PC^12,13,15–17^ with a limited range of prediction options whereas our study included patients with all stages (localized and metastasized) and all treatment lines of histologically confirmed UC and RCC. Thus, we have integrated a large number of patient and tumor characteristics, resulting in TR that contain the full spectrum of available anti-cancer treatments. “ChatGPT”^15^(® OpenAI) and “AI-Pathway Companion Prostate Cancer”^16,17^(® Siemens Healthineer) have been tested to support TR in localized PC based on current EAU- and National Comprehensive Cancer Network Guidelines with good results, but the studies included only 10 patient cases which lacks statistical power. Finally, data protection issues regarding sensitive patient data remain unsolved.

### New technical approaches in generating AI-based treatment recommendations

The main differentiation from existing methods is that different pre-existing approaches have been interactively adapted to the problem. The division of the TR into a two-stage system at different levels (High-level and Low-level) enables a recommendation based on a limited caseload.

Another unique technical feature is the associated multi-label prediction learning approach. Frequently, equivalent treatment options are recommended for a patient in MCC. This can only be achieved if the AI is able to predict multiple labels (multi-label prediction). To achieve this with a limited number of training cases, the classifier was modified by deliberately duplicating patient data with different labels. Either soft labelling or multi-label classifiers^18–20^ have been used in other studies, but both methods require a larger amount of data than the one used here.

In addition, our AI system includes a unique explainability component which allows the user to validate and, if necessary, optimize TR based on a transparent process. The challenge of explainability of machine prediction models has so far been addressed by various gradient-based, permutation-based or surrogate-based methods^21,22^. Based on the tabular patient data, SHAP^14^ proved to be the most suitable one due to its good visualization, running time and different data types (category, numeric, date).

The main methodological strength of this study is the manually extracted and quality-controlled patient data of 1429 (UC) and 786 (RC) original, retrospective MCC cases with corresponding 77 and 76 individual patient variables, respectively. This provides an unbiased approach to train the AI, avoiding pre-assumptions or pre-selection of patient factors that could potentially influence the subsequent recommendation. Another key advantage of our AI assistance system is its ability to be explained based on patient and clinical trial data. Explainability of AI is crucial in medical applications, as it facilitates user comprehension of the AI’s decision-making process. The main limitations of our study are the single-center and retrospective study design. In addition, some Low-Level features had to be summarized as “Other” due to small case numbers in this first step of AI development. Manual data extraction, currently considered the gold standard, may now be a limitation. However, we expect that suitable automated data extraction with comparable quality outcome will soon be available. Automated extraction was attempted and tested during the study, but was not yet feasible due to several limitations of the required use of LLM (insufficient quality, hallucinations, computer capacity).

The study represents a significant contribution to evidence-based multidisciplinary cancer care and demonstrates the feasibility of generating AI-based MCC recommendations of high clinical quality. The quality of the results relies on the time-consuming manual extraction process, which is currently mandatory due to the limitations of automated extractions, e.g. by LLM. Furthermore, the use of a two-stage prediction system facilitates the achievement of highly favorable outcomes, despite the limited number of cases.

### Conclusions

We present the first explainable AI support system for TR in UC and RCC. They are presented in a comprehensive dashboard including visualized explainability based on patient characteristics. This study pioneers future developments and sets a global standard for the development of AI clinical decision support routines in Clinical Oncology. Next, a prospective study will be conducted to provide a higher level of evidence and extend the AI system to prostate cancer. Once a successful foundation has been established in urological oncology, it would be possible to extend the AI system to other oncological diseases.

## Acknowledgments

We thank Yvonne Wiesner and Caecilia Dzalto (both University Medical Center Mainz) for their support in documenting patient data and Crispin Wiesmann (University Medical Center Mainz) for his support in documenting clinical trial data.

## Intellectual property

The AI developed in this study presented was registered for a European patent at the European Patent Office on 31 January 2025 (application number: EP25155356.6, title: system and method for assisting multidisciplinary decision-making for treatment recommendations).

## Data access and responsibility

All authors had full access to all the data in the study and responsibility for the integrity of the data and the accuracy of the data analysis.

## Informed consent and patient details

All procedures performed in studies involving human participants were in accordance with the ethical standards of the ethics committee of the Medical Association Rhineland-Palatinate, Germany (2022-16511, up-dated 2022-16511_2) and with the 1964 Helsinki Declaration and its amendments of ethical standards.

## Data Availability Statement

Currently, the original datasets are not available to other researchers due to the ethics committee of the Medical Association Rhineland-Palatinate, Germany (2022-16511, up-dated 2022-16511_2).

## Previous Presentations

Preliminary results were presented or will be presented at the following scientific meetings:

1. Duwe G, Mercier D, Kauth V, Moench K, Junker M, Vesga JP, Seiz W, Scheele J, Dengel A, Haferkamp A, Hoefner T. 013P First preliminary results of artificial intelligence generated treatment recommendations for urothelial cancer based on multidisciplinary cancer conferences from the KITTU project. 38th Annual European Society of Medical Oncology Congress, 13-17.09.2024, Barcelona. Annals of Oncology, Volume 35, Supplement 2, September 2024, Page S1161.
2. Duwe G, Mercier D, Kauth V, Moench K, Junker M, Dengel A, Haferkamp A, Hoefner T. First preliminary results of artificial intelligence-generated, explainable treatment recommendations for renal cell cancer based on multidisciplinary cancer conferences. American Society of Clinical Oncology Genitourinary Cancer Symposium, 13-15.02.2025, San Francisco, USA. Published only on 10th February 2025.

---

**Fig.S1:**
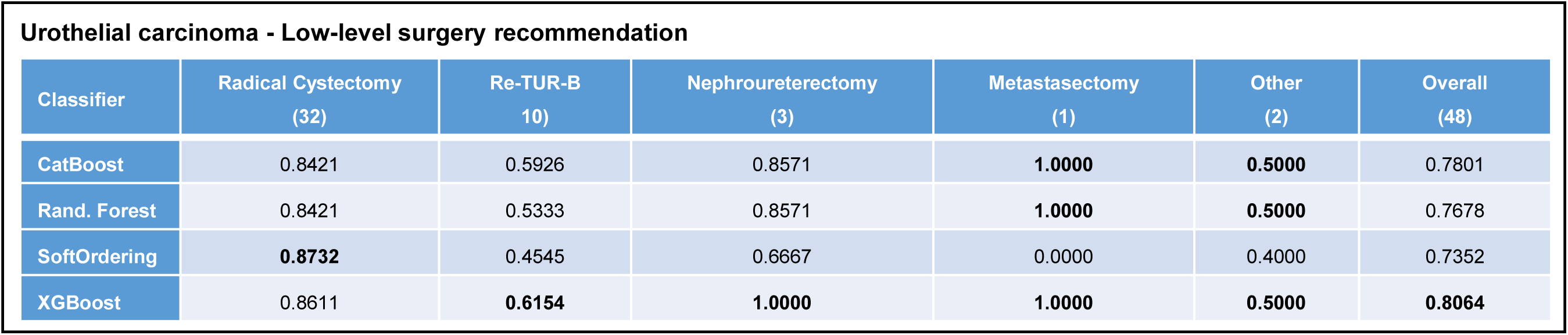
Urinary tract carcinoma: AI performance results (test set) of different classifiers for Low-level surgery TR. Numbers in brackets correspond to number of cases in the test set. Numbers in the table show the specific F1-scores. The ‘Other’ class always corresponds to a group of classes that is under-represented in the dataset and can be more specified in future once the dataset has a sufficient number of cases for a reasonable AI training.

**Fig.S2:**
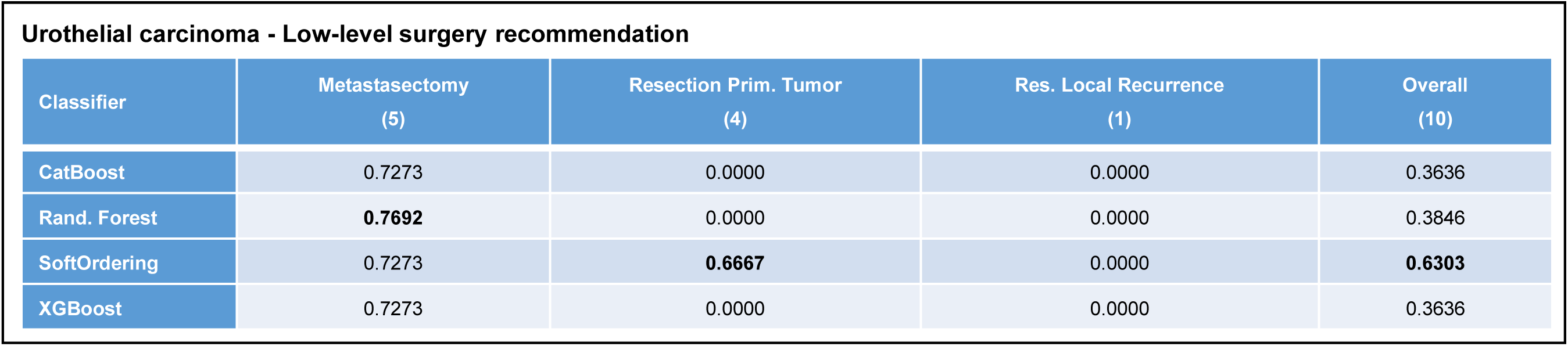
Renal cell carcinoma: AI performance results (test set) of different classifiers for Low-level surgery TR. Numbers in brackets correspond to number of cases in the test set. Numbers in the table show the specific F1-scores.

