## Supplement Material for "Development of an artificial intelligence-generated, explainable treatment recommendation system for urothelial carcinoma and renal cell carcinoma to support multidisciplinary cancer conferences"

### Supplementary Material

### S1. Materials and Methods

#### S1.1. Ethic statement and legal framework

The study was conducted in accordance with the Declaration of Helsinki and the International Ethical Guidelines for Biomedical Research Involving Human Subjects. The data protection officer of University Medical Center Mainz (UMCM) and the ethics committee of the Medical Association of Rhineland-Palatinate, Germany (2022-16511, up-dated 2022-16511_2) approved the study. Finally, the legal cooperation (including data protection) between UMCM and the German Research Center for Artificial Intelligence (DFKI) was agreed by means of a cooperation agreement and an order processing agreement.

#### S1.2. Data security

Patient data sets were pseudonymised by the Institute for Medical Biostatistics, Epidemiology and Informatics of UMCM. A randomly generated 10-digit number was used as pseudonym. Personal data (name, date of birth, patient-ID) was removed before further processing and can only be restored by assigned staff members of the study team at UMCM for quality check of results.

**S1.3. Data transfer between UMCM and German Research Center for Artificial Intelligence (DFKI)**

To ensure a secure transfer, a locally hosted cloud storage at DFKI with an authentication service was used to upload the pseudonymised data by UMCM. Afterwards, the data was downloaded from the cloud storage and processed (preprocessed and trained) by DFKI. After the final evaluation using the AI-generated treatment recommendation (TR) on the side of the storage, a result file is uploaded for the assessment by UMCM.

**S1.4. Data preprocessing and network training**

The downloaded data was technically adapted ("preprocessed") to be suitable for AI development. As the tabular data contains complex patient characteristics, it was mandatory to simplify the complex data types without any loss of information. Numerical values were normalized and standardized to ensure optimal training of the machine learning architecture. Categorical values were additionally converted to numerical values. For the complex data types such as the programmed death-ligand 1 scores, splitting into different components was performed using regular expressions, followed by a categorization step. A two-step process was developed to train a classifier to mimic MCC recommendations. Due to the dataset size which was relatively small for a machine learning approach and the fact that multiple, equal treatment recommendations are given in some cases, a special processing step has been deployed (confusion using duplicated patients) that makes the pipeline unique. Precisely, a counter intuitive decision was established for MCCs with multiple recommendations which were duplicated with their different recommendations (see Supplementary Material):

Since it was not possible to directly train a network for multi label prediction due to the low number of instances, a counter intuitive decision was established for MCCs with multiple equivalent recommendations by means of duplicating them to achieve single recommendations. It was ensured that such instances were always in the same (train or test) set. When a model is trained with duplicates, the distribution of prediction probabilities tends to be more evenly distributed rather than more predictive. A threshold can be used to determine the value at which a prediction is accepted as valid, making it feasible to specify multiple valid predictions with a single vector.

**S1.5. Description of the workflow of data processing (Fig.1)**

The upper blue part corresponds to the AI-generated TR including two different AI systems for the high-level and low-level classification. The bottom grey part applies to the implementation of clinical trial data to evaluate anti-cancer drug TR by applying two LLMs. The pipeline is separated into two different components each with multiple individual AI systems. The upper blue part handles the High-level and Low-level recommendation. For the High-level classification, a single AI is used, however, for the Low-level classification a separate AI is used for surgery and medication prediction. The lower, grey part is only performed if an 'Anti-cancer drug' has been recommended by KITTU. It shows the component that uses two AI systems to assess whether a clinical trial is applicable to a patient. This gives additional information and survival rates for the recommended and for comparable drugs, which aims to further enable the MCC physicians to modify the drug recommendation if a clinical trial suggests a more suitable drug.

#### S1.6. Explainability based on clinical trials of anti-cancer drugs

To enhance the recommendation and explainability, survival data of clinical trials for anti-cancer drugs were taken into account. Relevant clinical trials were identified from the current European Association of Urology (EAU) guidelines for muscle-invasive and metastatic bladder cancer, upper urinary tract urothelial cell carcinoma and renal cell cancer as well as from ClinicalTrials.gov. In- and exclusion criteria and survival data were manually extracted to an AI-readable table. Large Language Model (LLM) CapybaraHermes was used for analysis of clinical trial data and its correlation to KITTU patient data. The first LLM prompt extracts patient data relevant to the trial in- and exclusion criteria, the second LLM prompt checks whether the criteria are met.

### S2. Results – Supplementary information

#### S2.1. Performance results for urinary tract carcinoma - Low-level surgery recommendations

XGBoost achieved the best overall F1-score (0.8064) and the best performance for the classes nephroureterectomy (1.000) and metastasectomy (1.000) (Fig. S1). Regarding the low case numbers in the test set of only three nephroureterectomies and one metastasectomy, there is a certain possibility for a random hit of a correct recommendation. Since the F1-score of 1.000 describes the correctly recommended treatment in all four cases of nephroureterectomy and metastasectomy (100% sensitivity), it means at the same time that none of the other 44 cases with surgery recommendations have falsely recommended those two treatments (no false positive recommendations). This lowers the likelihood of merely random recommendations by the AI when facing low case numbers.

#### S2.2. Performance results for renal cell carcinoma - Low-level surgery recommendations

SoftOrdering showed the best overall F1 score of 0.6303. While the machine learning approach was able to learn to differ between the two classes ‘Metastasectomy’ (0.7273) and ‘Resection Primary Tumor’ (0.6667), it was not yet possible to sufficiently train the third class ‘Resection Local Recurrence’ due to the low number of cases. However, the prediction of the Metastasectomy produced very stable results in the test set.

#### S2.3. Explainability and Dashboard Presentation

The dashboard is divided into different components. The first component shows the High-level and Low-level TR. The automatic system can recommend multiple therapies which are equivalent and shows the probabilities for the recommendations. The next components address the explainability. First, the explanation of the individual TR is shown in a waterfall plot. The plot shows a subset of the features that affected the TR most. The last component (which is in the developing stage at the time of publication) will show the applicable clinical trial corresponding to the AI-recommended treatment based on its in- and exclusion criteria and its survival data. This LLM based AI-system matches trial inclusion criteria with the corresponding clinical parameters of the patient. Finally, further trials can be included in the visualization after checking their degree to which a patient can possibly apply to them. This will only be applicable for drug recommendations.

Particularly noteworthy is that the patient/clinical trial matching approach is using two LLMs, which splits the problem to make the prediction more reliable. The analysis and classification of clinical trial data and patient data by neural networks has not yet been tested. The comparison of patient data with clinical trial data in order to classify their applicability could be conditionally implemented using a rule set, but the approach would not be extensible (or only very cumbersome). In addition, the use of rule-based approaches is very limited, as the studies and patient data change over time, which makes constant, time-consuming adaptation essential. As an alternative, LLMs can be used, which are very flexible with regard to the data and can be easily adapted to new study criteria. It is also possible to have different representations, e.g. abbreviations or synonyms, evaluated by the AI. However, LLMs have the disadvantages of "hallucinating" and they have data protection problems (when using externally provided public networks)[29-31].

**Table S1. Clinical patient data structure:**

**Urinary tract carcinoma – 77 Input Features** (entry specified depending on patient data)

**General patient data**

- Age
- Sex
- ECOG Performance Status

Comorbidities

- Arterial hypertension
- Cardiovascular diseases
- Renal insufficiency
- Degree of renal insufficiency
- Dialysis
- Neurological diseases
- Malignancies (other than MCC diagnosis)

**Specific oncological data**

- Initial diagnosis of urinary tract carcinoma – date
- Initial diagnosis of urinary tract carcinoma – date known or estimated (k/e)

TMN Classification (histological)

- Localization
- Side (left or right, in case of affected kidney or ureter)

- Histological type
- T-stage (bladder cancer)
- T-stage (Ureteral or renal pelvis cancer)
- N-stage
- M-stage
- Grading
- R-classification
- Perineural invasion
- UICC
- Localization distant metastases
- PD-1 status
- PD-L1 status

Previous anti-cancer drug treatments (for MCC diagnosis)

- BCG (Bacillus Calmette-Guérin)
- Mitomycin
- Chemotherapy 1
- Number of cycles
- Chemotherapy 2
- Number of cycles
- Chemotherapy 3
- Number of cycles
- Chemotherapy 4
- Number of cycles
- ADC (antibody drug conjugate)
- Number of cycles

- Checkpoint inhibitor 1
- Number of cycles
- Checkpoint inhibitor 2
- Number of cycles
- Checkpoint inhibitor 3
- Number of cycles

Previous anti-cancer treatment (other than drugs)

- Transurethral resection of the bladder (TURB)
- Surgery
- Urinary diversion
- Radiotherapy
- Radiochemotherapy
- Other

Radiological imaging (most current staging, ≤3 months prior to MCC)

- Clinical T-stage
- Locoregional lymph node metastases
- Distant metastases
- Distant metastases localization

Laboratory values (most current analysis, ≤3 months prior to MCC)

- Leukocytes
- Hemoglobin
- Thrombocytes
- Neutrophil granulocytes
- GOT (ASAT)
- GPT (ALAT)

- gamma-GT
- Total bilirubin
- LDH
- Total protein
- Albumin
- Creatinine
- eGFR
- Sodium
- Potassium
- Calcium
- Urea-N
- Quick
- PSA

Current situation:

- Tumor progress / regress / stable disease
- Current UICC

MCC

- MCC date
- Upcoming treatment line

**Urinary tract carcinoma – 13 Output Features** (entry specified depending on patient data)

**MCC recommendation**

- Surgery
- Surgery (as additional equivalent recommendation)
- Radiotherapy
- Anti-cancer drug treatment
- Anti-cancer drug treatment (as additional equivalent recommendation )
- Anti-cancer drug treatment (as additional equivalent recommendation 2)
- Anti-cancer drug treatment (as additional equivalent recommendation 3)
- Continuation of current anti-cancer drug treatment
- Radio-chemotherapy
- Radio-chemotherapy (as additional equivalent recommendation)
- Follow-up
- Best supportive care
- Further diagnostics

**Renal cell carcinoma – 76 Input Features** (entry specified depending on patient data)

**General patient data**

- Age
- Sex
- ECOG Performance Status
- Karnofsky score

Comorbidities

- Arterial hypertension
- Cardiovascular diseases
- Thrombotic events
- Renal insufficiency
- Degree of renal insufficiency
- Dialysis
- Neurological diseases
- Metabolic disorders
- Malignancies (other than MCC diagnosis)
- Anti-cancer drug treatment for further malignancies

**Specific oncological data**

- Initial diagnosis of renal cell carcinoma – date
- Initial diagnosis of renal cell carcinoma – date known or estimated (k/e)
- MSKCC (Motzer) score
- IMDC prognostic score

TMN Classification (histological)

- Localisation (left or right kidney)
- Histological type
- T-stage
- N-stage
- M-stage
- Grading
- R-classification
- UICC
- Localization distant metastases 1
- Localization distant metastases 2
- Localization distant metastases 3

Previous anti-cancer drug treatments (for MCC diagnosis)

- Chemotherapy 1
- Number of cycles
- Chemotherapy 2
- Number of cycles
- Chemotherapy 3
- Number of cycles
- Chemotherapy 4
- Number of cycles
- Chemotherapy 5
- Number of cycles
- Chemotherapy 6
- Number of cycles

- Chemotherapy 7
- Number of cycles

Previous anti-cancer treatment (other than drugs)

- Radiotherapy
- Surgery
- Other

Radiological imaging (most current staging, ≤3 months prior to MCC)

- Clinical T-stage
- Locoregional lymph node metastases
- Distant metastases
- Distant metastases localization

Laboratory values (most current analysis, ≤3 months prior to MCC)

- Leukocytes
- Hemoglobin
- Thrombocytes
- Neutrophil granulocytes
- GOT (ASAT)
- GPT (ALAT)
- gamma-GT
- Total bilirubin
- LDH
- Total protein
- Albumin
- Creatinine
- eGFR

- Sodium
- Potassium
- Calcium
- Urea-N
- Quick
- PSA

Current situation

- Tumor progress
- Tumor regress
- Stable disease
- Mixed response
- Current UICC

MCC

- MCC date
- Upcoming treatment line

**Renal cell carcinoma - 11 Output Features** (entry specified depending on patient data)

**MMC recommendation:**

- Surgery
- Surgery (as additional equivalent recommendation)
- Radiotherapy
- Anti-cancer drug treatment
- Anti-cancer drug treatment (as additional equivalent recommendation)
- Anti-cancer drug treatment (as additional equivalent recommendation 2)
- Anti-cancer drug treatment (as additional equivalent recommendation 3)
- Anti-cancer drug treatment (as additional equivalent recommendation 4)
- Continuation of current anti-cancer drug treatment
- Follow-up
- Best supportive care
